# Knockdown resistance (kdr) Associated organochlorine Resistance in mosquito-borne diseases (*Culex pipiens*): Systematic study of reviews and meta-analysis

**DOI:** 10.1101/2023.12.17.23300098

**Authors:** Ebrahim Abbasi, Salman Daliri, Asghar Talbalaghi, Fatemeh Mehrpouya, Maryam Hasanzadeh arab, Atena Aslvaeli

**Affiliations:** Research Center for Health Sciences, Institute of Health, Shiraz University of Medical Sciences, Shiraz, Iran; Department of Medical Entomology and Vector Control, School of Health, Shiraz University of Medical Sciences, Shiraz, Iran; Research Clinical Research Development Unit, Imam Hossein Hospital, Shahroud University of Medical Sciences, Shahroud, Iran; italian Mosquito Control Association; Department of Medical Entomology, Faculty of Health, Ahvaz Jundishapur University of Medical Sciences, Iran

**Keywords:** Knockdown resistance, Organochlorine insecticide, *Culex pipiens*, Systematic study of reviews and meta-analysis

## Abstract

**Background:** *Culex pipiens* is the vector of a large number of pathogenic pathogens in humans. Using insecticides to deal with this vector is the most important way to control it. However, in recent decades, resistance to insecticides has been reported in this vector. One of the main insecticides used to fight this vector is organochlorine insecticides. Based on this, this study was conducted to investigate the prevalence of Knockdown resistance (kdr) in *Culex pipiens* against organochlorine insecticides.

**Methods:** This study was conducted by systematic review and meta-analysis in the field of kdr prevalence in *Culex pipiens* against organochlorine insecticides. Based on this, during the search in the scientific databases PubMed, Web of Science, biooan.org, Embase, ProQuest, Scopus, and Google Scholar without time limit until the end of November 2023, all related articles were extracted and analyzed. The statistical analysis of the data was performed using the random and fixed effects model in the meta-analysis, Cochran’s test, *I^2^* index, and meta-regression by STATA software version 17.

**Results:** seven studies with a sample size of 2,029 *Culex pipiens* were included in the meta-analysis process. Based on the findings, the kdr resistance prevalence against Deltamethrin, Malathion, Permethrin, and DDT insecticides was estimated as 30.6%, 42%, 17.9%, and 76.3% respectively. Among them, the highest resistance to DDT and the lowest to Permethrin was observed.

**Conclusion:** Based on the findings, a large proportion of *Culex pipiens* mosquitoes were resistant to DDT insecticide. However, this vector was highly sensitive to Deltamethrin, Malathion, and Permethrin insecticides. Due to the different resistance ratios in different regions of the world, it is recommended to conduct studies on the prevalence of kdr in *Culex pipiens*.

## Introduction

Culex is one of the most widespread mosquito species in the world (1). They opportunistically feed on humans and animals, this way of feeding provides suitable conditions for the transmission of common diseases between humans and animals and is a serious threat to public health (2). Culex has adapted to human habitats over the years and are expanding in the urban environment due to the rapid and unplanned expansion of cities and the lack of suitable environmental conditions, the presence of stagnant water, drains, organically polluted places, and pits (3, 4). *Culex pipiens* is one of the most important groups of Culex, which includes six members of *Cx. pallens Coquillet, Cx. Quinquefasciatus Say, Cx. australicus Dobrotworsky & Drummond, Cx. molestus Forskell, Cx. pipiens Linneaus and Cx. Globocoxitus* is Dobrotworsky (5, 6). *Culex pipiens* is an important vector of a large number of pathogenic pathogens and parasites in the world. This mosquito is known as a vector of West Nile virus (WNV), Rift Valley fever virus (RVFV), *Wuchereria bancrofti* (Cobbald, the causative agent of filariasis), and Japanese and St. Louis encephalitis (7–9).

Diseases transmitted through vectors (mosquitoes) continue to affect the public health of humans. Due to the lack of vaccination to prevent vector-borne diseases, combating them is considered the best method of intervention. As a result, nowadays chemical insecticides are mainly used to control the vectors. Four groups of insecticides: organochlorines, organophosphates, carbamates, and pyrethroids are the main insecticides used to deal with vectors (10). Pyrethroids account for about 15% of the insecticides used to combat vectors in the world. Pyrethroids are widely used to control vectors due to their low toxicity to humans and high killing effect on insects (11). Organochlorines have also been widely used since the distant past to fight against vectors. By acting on the central and peripheral nervous system of insects, these insecticides lead to paralysis and death of carriers through interaction with the voltage-gated sodium channel (VGSC) and increasing its sensitivity to depolarization by inhibiting inactivation processes (12, 13). As a result of the long-term, extensive, and incomplete use of these insecticides over time, it has led to the natural selection of insects and a reduction in the sensitivity of the target site to them, which is known as “KDR”. which is caused by the insensitivity of the target site due to the mutation in the voltage-sensitive sodium channel gene (Vssc) (14). Resistance to DDT and deltamethrin is often associated with mutations in the sodium channel gene, which reduces neuronal sensitivity to these insecticides (15). In the studies conducted in the world, kdr resistance was reported in *Culex pipiens* and it was shown that the ratio of resistance is different in different countries (16). Studies have mentioned that three groups of glutathione-S-trans-ferases (GST), esterase, and cytochrome P450 oxidases play a role in creating metabolic resistance to organochlorine, organophosphate, and pyrethroids in *Culex pipiens* (17). In the field of Kdr resistance in *Culex pipiens*, it has been reported that two mutations L1014F and L1014S cause Kdr resistance in it (18).

Identifying mutations related to resistance is essential for managing and using appropriate and effective insecticides to control insects. Considering that *Culex pipiens* is a carrier of some pathogenic pathogens for humans and is now spreading in the world (19, 20). Determining the level of sensitivity or resistance to insecticides is essential to deal with this vector. Based on this, the present study conducted to investigate the prevalence of kdr resistance in *Culex pipiens* against organochlorine insecticides via a systematic review and meta-analysis.

## Material and Methods

This study was conducted by systematic review and meta-analysis based on the guidelines of the Preferred Reporting Items for Systematic Reviews and Meta-Analyses (PRISMA) in the field of kdr prevalence in *Culex pipiens* against organochlorine insecticides (21). This research has been registered in the International Prospective Register of Systematic Review (PROSPERO) with the code CRD42021231605.

### Search Strategy

Articles by two researchers in scientific databases PubMed, Web of Science, biooan.org, Embase, ProQuest, Scopus, and Google Scholar using keywords Resistance, knockdown resistance, KDR, insecticide, Organochlorine insecticide, chlorinated insecticide, chlorophenyl, dichloroethane, DDT, parachlorophenyl, dichlorodiphenyldichloroethane, dieldrin, permethrin, deltamethrin, malathion, Culex and *Culex pipiens* extract and investigate in the title, abstract and full text of the articles singularly and in combination using OR, AND and NOT operators without time limit until the end of November 2023.

### Inclusion and Exclusion Criteria

Based on the PICO model, published English-language articles conducted on *Culex pipiens* investigated kdr resistance to organochlorine insecticides, and the prevalence of resistance or mortality in exposure to organochlorine insecticides. were reported and had good quality and were included in the study. The articles that were conducted on other insects, and other insecticides (except organochlorine insecticides) were investigated in them, kdr resistance was not investigated in them, they lacked the desired quality, and in the review method, case report or Letters to the editor were excluded from the study.

### Quality Assessment

The quality assessment of the articles was done based on 22 parts of the STROBE (Strengthening the Reporting of Observational Studies in Epidemiology) checklist, which investigated compliance with the principles of writing and implementation in the title, the method of reporting findings, limitations, and conclusions. Each part of this checklist is given a score based on its importance; the maximum possible score is 33. Based on the obtained score, the studies were divided into three levels low, medium, and high quality (22).

### Data Extraction

First, the articles were investigated by two researchers independently by investigating the title and abstract, taking into account the inclusion and exclusion criteria. Then, the full text of the articles was investigated by these researchers, and if the articles were rejected by two researchers, the reason was mentioned, and in case of disagreement between them, the article was refereed by a third researcher. Data extraction was done using a pre-prepared checklist that included the first author’s name, study place, study year, sample size, insecticide type, kdr resistance prevalence, and mortality.

### Selection of Studies

The number of 14536 studies were extracted by searching the databases. At first, the articles were entered into the Endnote software, and after the initial review, 6528 articles were excluded from the study due to duplicates. Then, by checking the titles and abstract of the articles, 7839 articles were removed because they were not relevant, and after reviewing the full text of the articles, 162 articles were removed due to the lack of investigation of the prevalence of kdr resistance or resistance to organochlorine insecticide, and 7 articles met the inclusion criteria. and entered the meta-analysis process (Figure 1).

**Figure 1.**
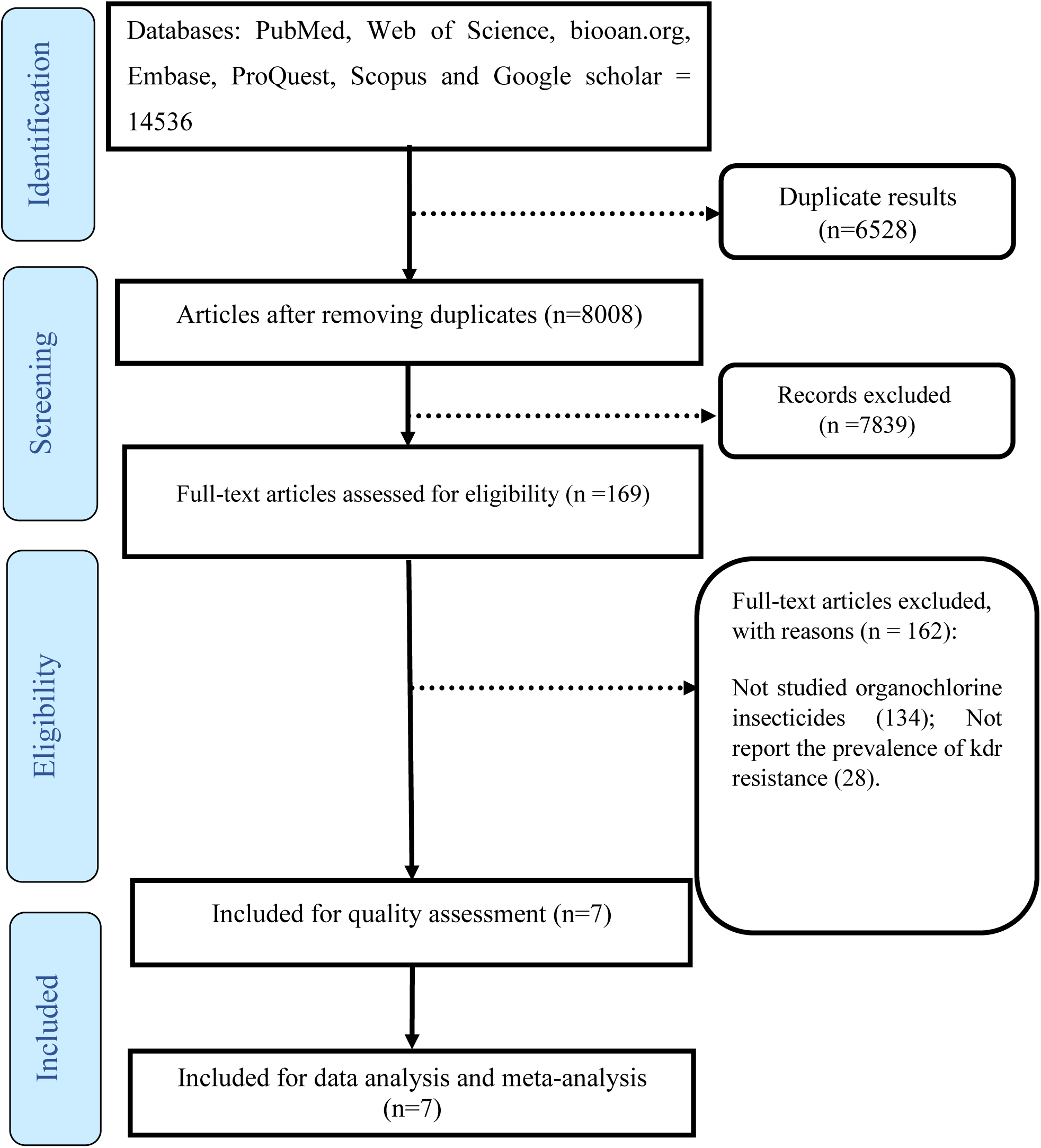
The PRISMA flow diagram

### Statistical Analysis

Statistical analysis of the data was done using random and fixed effects models in meta-analysis, *I^2^* index, and Cochran’s test. funnel plot was used to investigate of the publication bias and Meta regression to investigate the relationship between the sample size and the prevalence of KDR. Data analysis was done using STATA software version 17.

## Results

Seven studies with a sample size of 2029 *Culex pipiens* mosquitoes that were conducted between 2006 and 2023 were included in the study. Two studies were conducted in China, two studies in Iran, and one study each in Morocco, Egypt, and America. The characteristic of the reviewed articles is presented in Table 1.

**Table 1.**
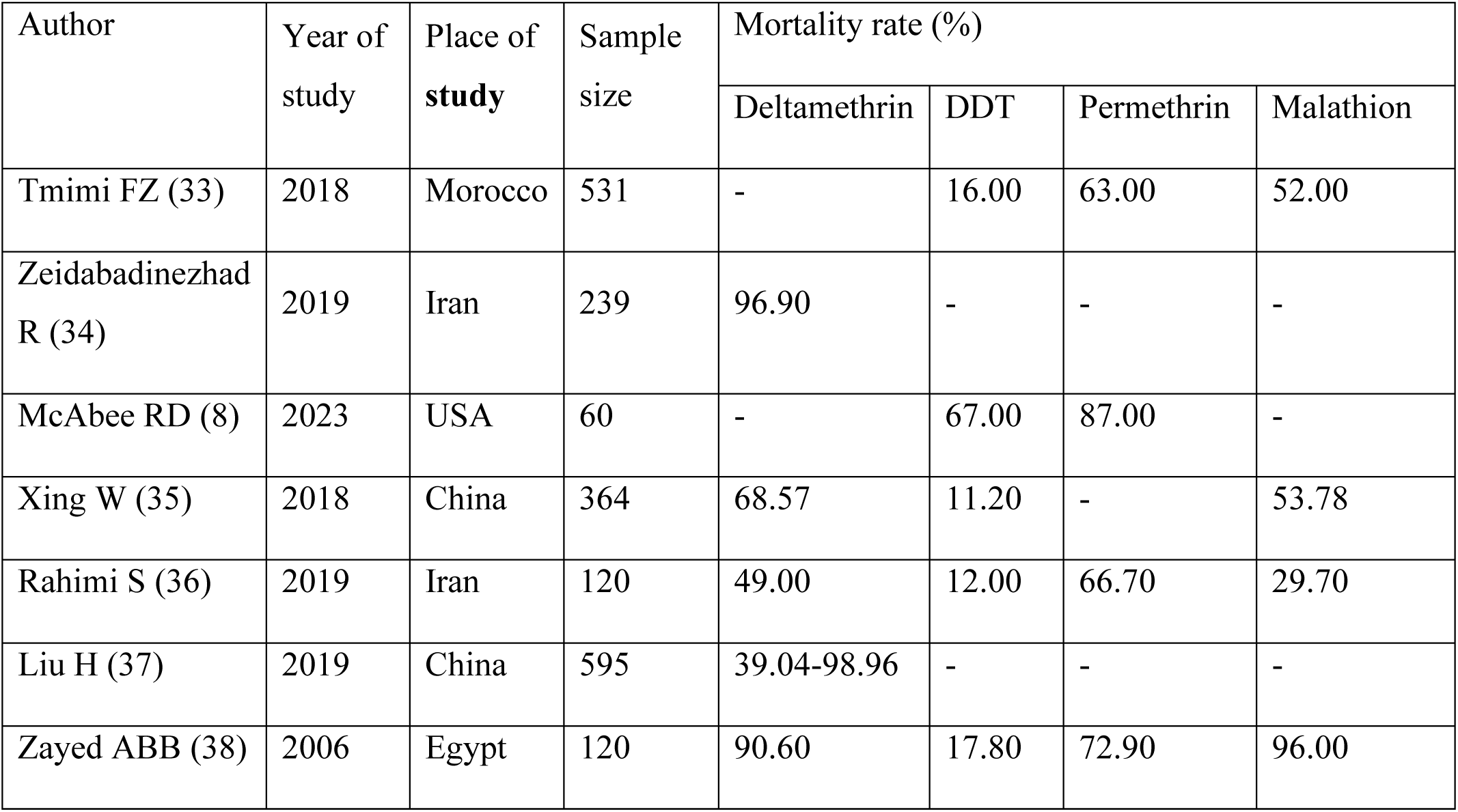
characteristic of the articles included in the meta-analysis.

| Author | Year of study | Place of study | Sample size | Mortality rate (%) |  |  |  |
| --- | --- | --- | --- | --- | --- | --- | --- |
|  |  |  |  | Deltamethrin | DDT | Permethrin | Malathion |
| Tmimi FZ (33) | 2018 | Morocco | 531 | - | 16.00 | 63.00 | 52.00 |
| Zeidabadinezhad R (34) | 2019 | Iran | 239 | 96.90 | - | - | - |
| McAbee RD (8) | 2023 | USA | 60 | - | 67.00 | 87.00 | - |
| Xing W (35) | 2018 | China | 364 | 68.57 | 11.20 | - | 53.78 |
| Rahimi S (36) | 2019 | Iran | 120 | 49.00 | 12.00 | 66.70 | 29.70 |
| Liu H (37) | 2019 | China | 595 | 39.04-98.96 | - | - | - |
| Zayed ABB (38) | 2006 | Egypt | 120 | 90.60 | 17.80 | 72.90 | 96.00 |

Based on the findings of the meta-analysis of six studies conducted on the prevalence of kdr resistance in *Culex pipiens* against Deltamethrin insecticide, the ratio of sensitivity to Deltamethrin insecticide was estimated at 69.4%. which shows that 30.6% of *Culex pipiens* mosquitoes were resistant to Deltamethrin. In terms of heterogeneity between studies, the *I^2^*index was estimated at 98.41%, which indicates the existence of heterogeneity between studies (Figure 2). The investigation of the prevalence of *Culex pipiens* resistance against Malathion insecticide showed that the prevalence of sensitivity in *Culex pipiens* mosquitoes was 58.1%, which indicates that about 42% of *Culex pipiens* have kdr resistance to Malathion (Figure 3).

**Figure 2.**
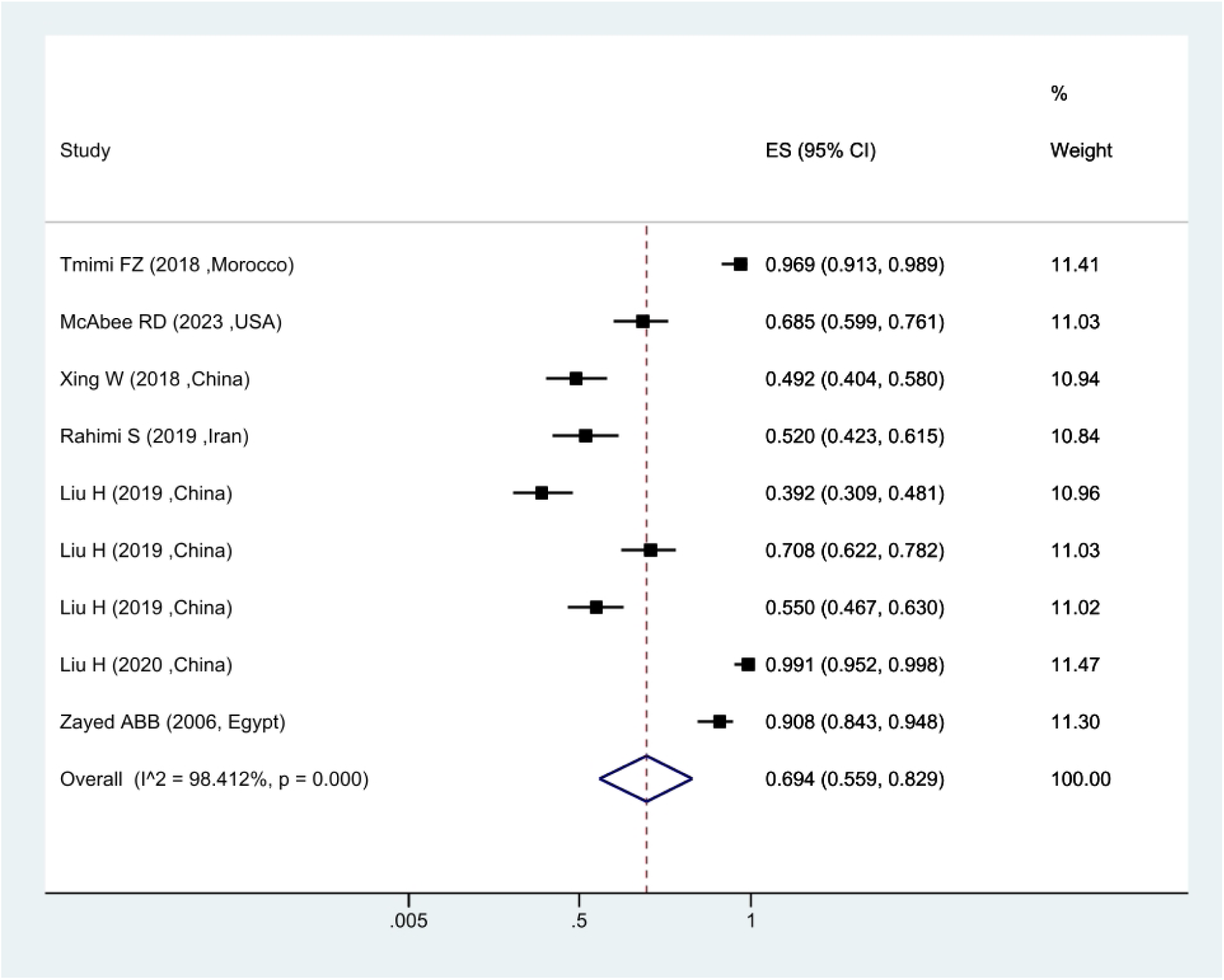
Forest plots of the mortality rate *Culex pipiens* exposed to Deltamethrin and 95% confidence interval based on the random effect model in meta-analysis.

**Figure 3.**
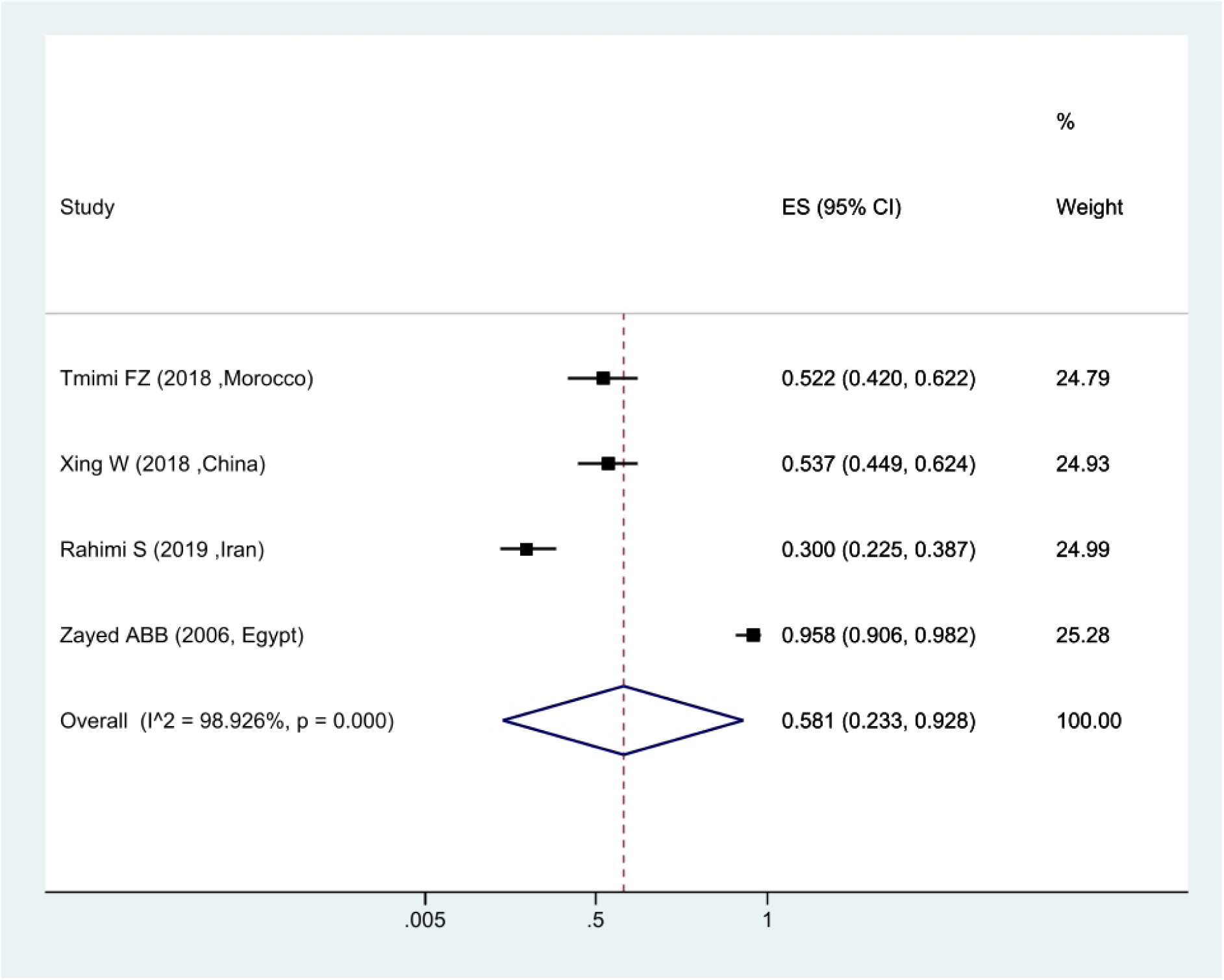
Forest plots of mortality rate *Culex pipiens* exposed to Malathion and 95% confidence interval based on the random effect model in meta-analysis.

In the context of the prevalence of kdr resistance in *Culex pipiens* against Permethrin insecticide, the findings showed that 72.1% of *Culex pipiens* were sensitive to Permethrin and it shows that 17.9% of them have kdr resistance (Figure 4). The attenuation ratio against DDT insecticide was estimated at 23.7%, which shows that 76.3% of *Culex pipiens* have kdr resistance against DDT (Figure 5).

**Figure 4.**
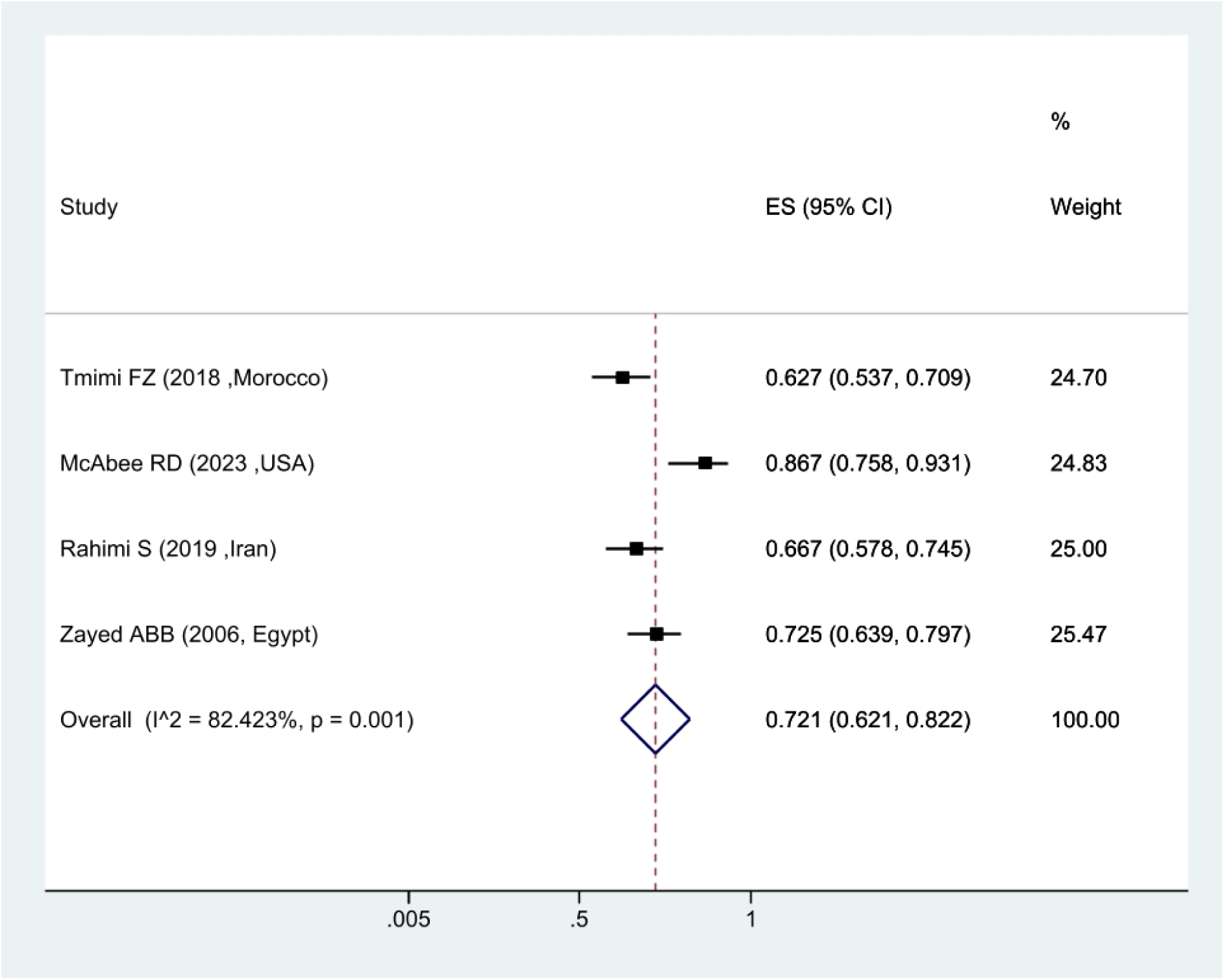
Forest plots of mortality rate *Culex pipiens* exposed to Permethrin and 95% confidence interval based on the random effect model in meta-analysis.

**Figure 5.**
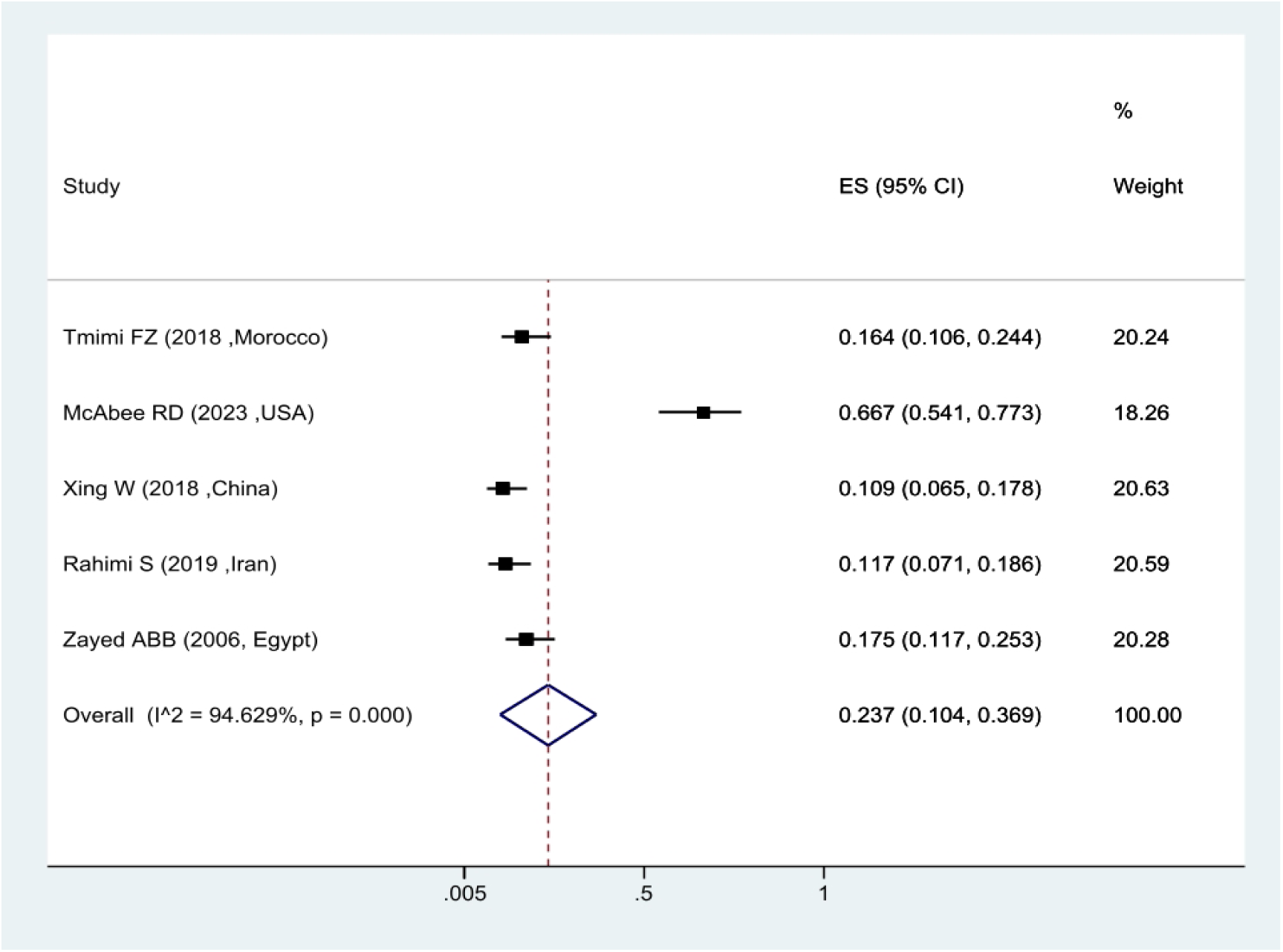
Forest plots of mortality rate *Culex pipiens* exposed to DDT and 95% confidence interval based on the random effect model in meta-analysis.

The publication bias was done using the funnel plot and Egger’s test. Due to the symmetry of the funnel plot, it can be mentioned that publication bias did not occur, and the result of Egger’s test was not significant in this regard (P=0.14) (Figure 6). Also, using meta-regression, the relationship between sample size and mortality ratio was investigated. According to the slope of the graph, the mortality ratio decreased with the increase of the sample size. which shows that the resistance ratio is higher in larger populations (Figure 7).

**Figure 6.**
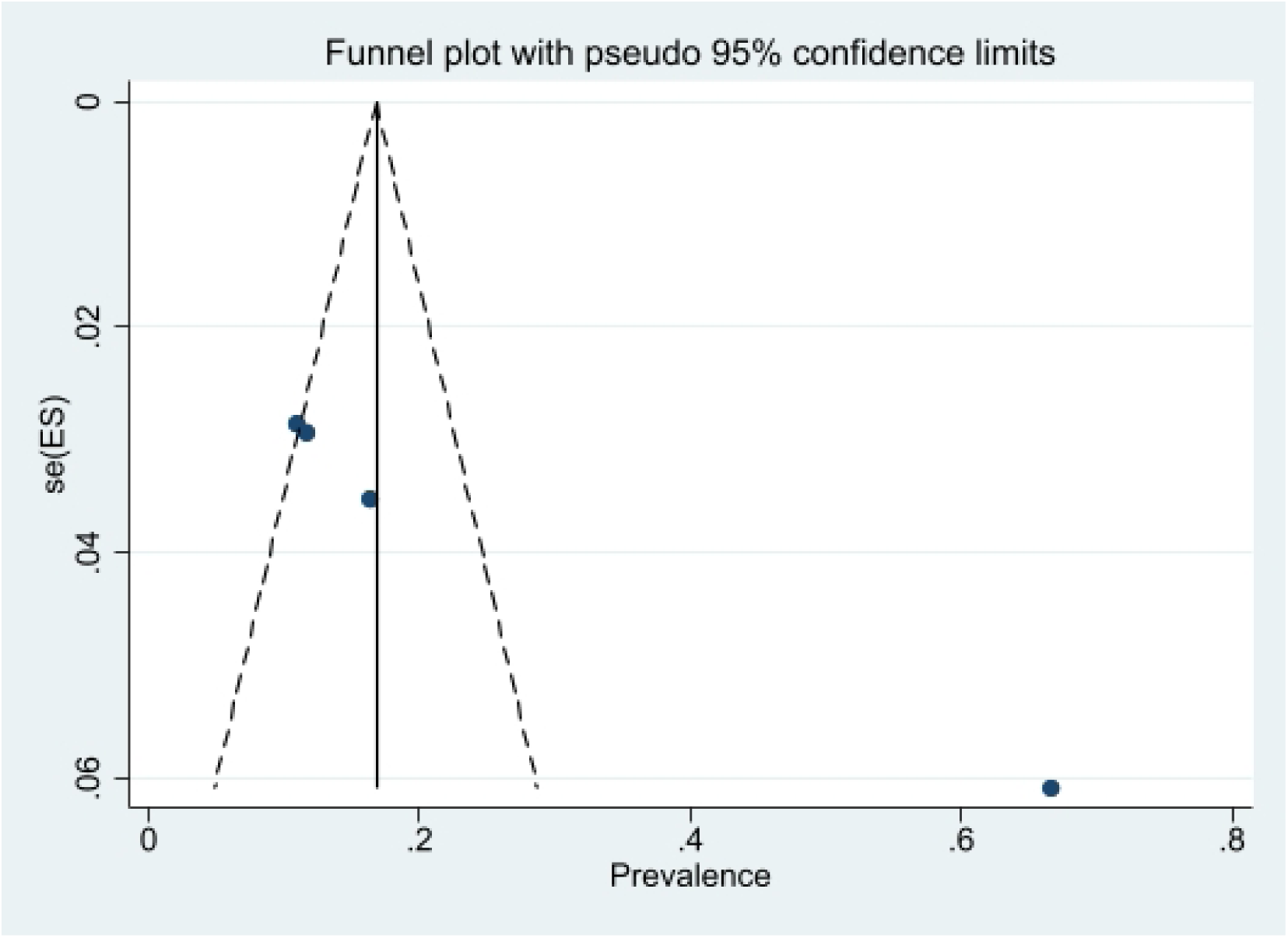
Funnel plot of the mortality rate *Culex pipiens* in the selected studies

**Figure 7.**
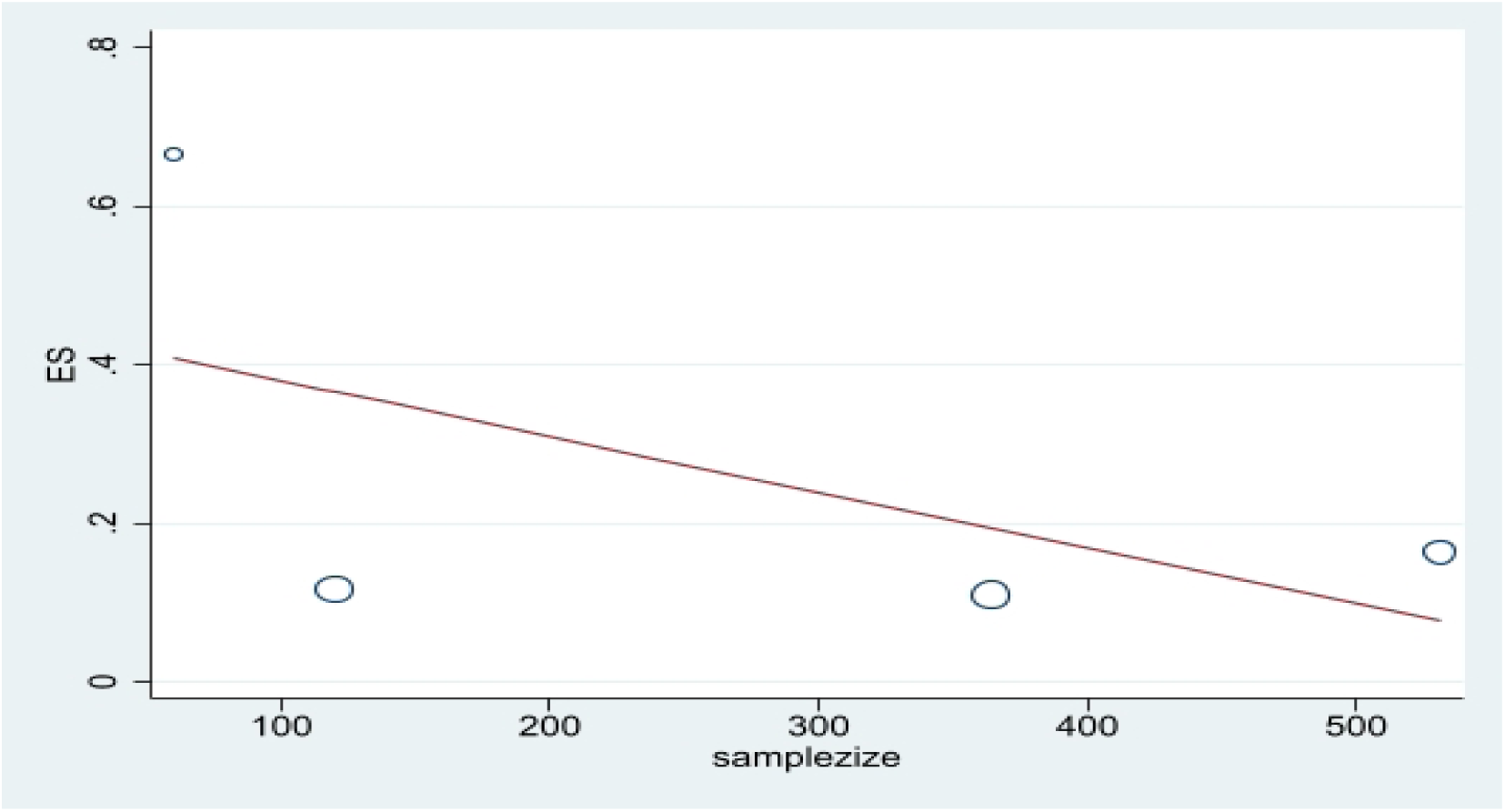
Meta regression plot of the mortality rate *Culex pipiens* of exposed to Permethrin based the study year.

## Discussion

This study was conducted on the prevalence of kdr resistance in *Culex pipiens* against Deltamethrin, Permethrin, Malathion, and DDT insecticides by meta-analysis method. Based on the findings, the kdr resistance ratio against Deltamethrin, Malathion, Permethrin, and DDT insecticides was estimated as 30.6%, 42%, 17.9%, and 76.3% respectively. Among them, the highest resistance to DDT and the lowest to Permethrin was observed. It can be mentioned that the most effective insecticide to deal with *Culex pipiens* is Permethrin and Deltamethrin. In studies of insecticide target sites in *Culex pipiens*, G119S ace-1 and L1014F kdr mutants. Regarding resistance to DDT, these mutations were identified and shown to play a role in creating resistance in *Culex pipiens* (23, 24). Studies have also shown that the L1014F kdr mutation is widely present in *Culex pipiens*. that this mutation plays an important role in resistance to organochlorine and organophosphate insecticides (25). The mutation from leucine to serine (TTA to TCA) is another mutation identified in *Culex pipiens* resistance, which has been observed in different countries including China, America, and Japan (26–28). The L1014C mutation with the substitution of leucine (TTA) for cysteine (TGT) is another mutation identified in *Culex pipiens* (26). Mosquitoes with the Phe/kdr mutation also showed high resistance to pyrethroids and DDT, but the Ser/kdr mutation confers high resistance to DDT and low resistance to pyrethroids (29). The presence of various mutations in *Culex pipiens* indicates the presence of high resistance and their spread in different regions of the world. Because under selection pressure, carriers can acquire resistance and transfer it to the next generations, as a result, this phenomenon can lead to an increase in the prevalence of kdr resistance in *Culex pipiens* mosquitoes in the world. In a study conducted in northern Iran, the sensitivity ratio of *Culex pipiens* mosquitoes to DDT was low, but a high ratio of them was sensitive to deltamethrin (30). In another study by Salim-Abadi et al. (2016) in Iran, *Culex pipiens* were sensitive to deltamethrin and resistant to DDT (31). In the study of Akiner et al. (2009) in Turkey, *Culex pipiens* was highly sensitive to malathion, deltamethrin, and permethrin insecticides, but highly resistant to DDT (32).

In general, based on the findings of the present study, it can be mentioned that *Culex pipiens* is highly sensitive to Deltamethrin, Malathion, and Permethrin insecticides, but it has high KDR resistance to DDT insecticide. Based on this, it is recommended to use effective insecticides to fight and control this vector. Also, due to the different resistance ratios in different regions of the world, it is recommended to conduct studies on the prevalence ratio of kdr resistance.

## Conclusion

According to the findings, a large proportion of *Culex pipiens* mosquitoes were resistant to DDT insecticide. However, this vector was highly sensitive to Deltamethrin, Malathion, and Permethrin insecticides. Considering that few studies have been done in this field. It is recommended to conduct studies to evaluate the prevalence of resistance in countries where this vector is endemic.

## Declaration

### Ethical considerations

Ethical approval for this study was obtained from the Shiraz University of Medical Sciences (Iran) Ethics Committee.

### Availability of data and materials

All obtained data are included in the text.

### Authors’ contributions

EA determined the title, wrote and registered the protocol, and submitted the article. EA and FM extracted the files from the databases. AT, MH, and EA, screening, and selection of final articles. AA and EA, data extraction. SD wrote the article. All authors read and approved the final manuscript.

## Data Availability

All data produced in the present work are contained in the manuscript

## Acknowledgments

The authors thank the Research Vice-Chancellor of Shiraz University of Medical Sciences.

